# The prevalence, incidence and risk factors of mental health problems and mental health services use before and 9 months after the COVID-19 outbreak among the general Dutch population. A 3-wave prospective study

**DOI:** 10.1101/2021.02.27.21251952

**Authors:** Peter G. van der Velden, Miquelle Marchand, Marcel Das, Ruud Muffels, Mark Bosmans

## Abstract

**Objectives:** Gain insight in the effects of the COVID-19 pandemic on the prevalence, the incidence and risk factors of mental health problems among the Dutch general population and different age groups in November-December 2020, compared to the prevalence, incidence, and risk factors in the same period in 2018 and 2019. More specifically, the prevalence, incidence and risk factors of anxiety and depression symptoms, sleep problems, fatigue, disabilities due to health problems, use of medicines for sleep problems, medicines for anxiety and depression, and mental health services use.

**Methods:** We extracted data from the LISS (Longitudinal Internet studies for the Social Sciences) panel that is based a probability sample of the Dutch population of 16 years and older by Statistics Netherlands. We used three waves of the longitudinal Health module held in November-December 2018 (T1), November-December 2019 (T2) and November-December 2020 (T3), and selected respondents who were 18 years and older at T1. Data were weighted using 16 demographics profiles of the Dutch adult population (N^study sample^=4,064). The course of mental health problems was examined with repeated measures multivariate logistic regression analyses, and the differences in incidence with multivariate logistic regression analyses. In both types of analyses, we controlled for sex, age, marital status, employment status, education level and physical disease.

**Results:** The repeated measures multivariate logistic regression analyses among the total study sample did not reveal a significant increase in the prevalence of anxiety and depression symptoms, sleep problems, fatigue, disabilities due to health problems, use of medicines for sleep problems, medicines for anxiety and depression, and mental health services use in November-December 2020, compared to November-December 2018 and 2019 (that is, T3 did not differ from T1 *and* T2). Among the four different age categories (18-34, 35-49, 50-64, and 65 years old and older respondents), 50-64 years respondents had a significantly lower prevalence of anxiety and depression symptoms at T3 than at T1 and T2, while T1 and T2 did not differ. A similar pattern among 65+ respondents was found for mental health services use. We found no indications that the incidence of examined health problems at T2 (no problem at T1, problem at T2) and T3 (no problem at T2, problem at T3) differed. Risk factors for mental health problems at T2 were mostly similar to risk factors at T3; sex and age were less associated with sleep problems at T3, compared to T2 .

**Conclusions:** The prevalence, incidence and risk factors of examined mental health problems examined nine months after the COVID-19 outbreak appear to be very stable across the end of 2018, 2019 and 2020 among the Dutch adult population and different age categories, suggesting that the Dutch adult population (20 years and older) in general is rather resilient given all disruptions due to this pandemic.

## Introduction

The ongoing COVID-pandemic and preventive measures to contain the pandemic as much as possible have profound negative effects on affected countries and their residents. These effects vary from, but are not restricted to, higher death rates, recovery problems among infected, overloaded hospitals, closed schools, and diminished social contacts, to job loss, repeated lockdowns, political tensions, diminished economic growth, and large governmental financial debt. The global weekly Operational Update on COVID-19 of the World Health Organization of November 6 2020 and December 21 2020 [1], the period in which the last survey of the present study was conducted, reported 1,231,017 and 1,690,061 deaths respectively and 48,534,508 and 75,704,857 confirmed COVID-19 cases respectively.

Although effective vaccines have recently become available [2], it is unclear when this pandemic will end and related problems eventually will reach pre-COVID-19 outbreak levels or disappear. The question to what extent this ongoing pandemic affect the mental health of the general population is and remains therefore important. People, according to the Conservation of Resources (COR) model [3,4] strive to obtain, retain, protect resources and to restore lost resources (such as social contacts, employment, housing, health) and their resilience should not be underestimated [3-6]. The duration of this pandemic may, however, undermine the capacities of individuals, communities, and countries to cope with the negative effects of this pandemic on the medium and longer term.

To address this key question accurately, prospective studies based on probability samples of the general populations with pre-COVID-19 data on mental health are warranted, preferably with assessments at different time points after the outbreak because of the enduring character of this pandemic. Fortunately, an increasing number of peer-reviewed prospective population based-studies with pre-COVID data on mental health have been published. However, compared to the large number of cross-sectional COVID-19 studies that are often based on convenience samples for which the representativeness is unclear [7], the number of prospective studies based on probability samples is still very limited. In the meta-analysis by Santabárbara et al. [8] of 43 identified cross-sectional studies (until August 23, 2020) based on community samples, no study was based a on probability sample of the general population.

Being aware that (almost) accepted but not published peer-reviewed studies are missed, to date we identified 12 studies (written in English) among the general population in the United Kingdom (UK), United States of America (USA) and the Netherlands that were based on probability samples of the general population with non-retrospective data on pre-COVID-19 mental health. Below, we first provide a summary of the main outcomes of these studies. Given the aim of the present study (see below), we focus on the prevalence of mental health problems among the total study samples and different age categories.

## Results prospective studies UK

Identified studies among the general UK population are all based on the *UK Household Longitudinal Study*. Main findings of the study by Pierce et al. [9] showed that mean scores on the GHQ-12 increased significantly from 11.5 in 2018–2019 (data were collected year-round) to 12.6 in April 2020 (based on an extra COVID-19 web survey with, according to the authors, a lower response rate than normal). This increase was not considered a simple continuation of previous upwards trends from 2014 to 2019. The prevalence of clinically significant mental distress according to the GHQ-12 increased from 18.9% in 2018–2019 to 27.3% in April 2020. The increase in mental distress appeared to be largest in respondents 18–24 and 25–34 years old.

The study by Proto and Quintana-Domeque [10] showed similar findings up to April 2020 but focused especially on ethnicity and gender. Niedzwiedz et al. [11] also examined smoking, drinking and the separate items of the GHQ-12 up to April 2020. Their results showed that smoking declined (Relative Risk (RR)=0.9) and that the proportion of people drinking four or more times per week increased (RR=1.4) as did binge drinking (RR=1.5). Analyses of separate GHQ item scores showed especially a relatively large increase in being less able to enjoy day-to-day activities (about 17% to about 41%). Daly et al. [12] analyzed data on mental health up to June 2020. Their results showed that the prevalence of mental health problems according to the GHQ-12 increased from 24.3% in 2017–2019 to 37.8% in April, 34.7% in May, and 31.9% in June 2020. Although elevated in June 2020 compared to 2017-2019, the prevalence was lower (- 5.9%) than in April 2020. Respondents of 18-34 years old showed the largest increase in the prevalence of mental health problems (18.6%) until April 2020, but also the largest decline (- 9.8%) in problems between April and June 2020.

## Results prospective studies USA

Prospective studies in the USA are, in contrast to the UK, all based on different study samples. Twenge and Joiner [13] compared anxiety and depression symptomatology (according to the PHQ-2 and GAD-2) among adults in the *National Health Interview Survey* (NHIS; January-June 2019) with adults in the *Household Pulse Survey* (HPS; April-May 2020). Results showed that respondents in the HPS were three to four times more likely to screen positive for anxiety or depression disorders, or screen positive for both in April-May 2020, compared to respondents in the NHIS. About 30% screened positive for these disorders during the pandemic. Importantly, the NHIS participants were asked about symptoms in the last two weeks and HPS participants were asked about symptoms in the last seven days. McGinty et al. [14] also compared the mental health of adults before and after the outbreak of two different samples, e.g., respondents in the NHIS in 2018 and the *Johns Hopkins COVID-19 Civic Life and Public Health Surve*y in April 2020. According to the Kessler’s K6 Scale, in the first sample (2018) serious psychological distress was significantly lower (3.9%) than in the second sample (13.6%, April 2020). In April 2020, symptoms of psychological distress were highest among young adults aged 18 to 29 years (24.0% versus about 4% in 2018). Daly et al. [15] compared the prevalence of depression symptoms according to the PHQ-2 among adults of the *National Health and Nutrition Examination Survey* (NHNE) of 2017-2018, and adults in the *Understanding America Study* (UAS) in March and April 2020. Results showed a significant higher prevalence in March (10.6%) and April 2020 (14.4%) in the UAS samples compared to the 2017-2018 NHNE sample (8.7%). Among 18-34 years old respondents the increase of the prevalence of depression symptom was 7.3% between pre-COVID and March 2020, and 13.4% between March and April 2020. Among 35-54 years old respondents and respondents of 65 years and older, a significant but smaller increase between March and April 2020 was found (4.5% and 3.5% respectively). Breslau et al. [16] examined psychological distress according to the Kessler’s K6 Scale among respondents of the *RAND American Life Panel* (ALP) of adults age 20 and over. Results showed that the prevalence of clinical psychological distress in February 2019 (10.9%) was as high as in May 2020 (10.2%). Importantly, in this study the K6 questions in February 2019 referred to the worst month of the past year (before February 2019) while the May 2020 assessment referred to the past-30 days. Among 20-39 and 40-59 years old respondents the prevalence of respondents who suffered from an increase of problems (20.8% and 14.4% respectively) was higher than among those of 60 years and older. Ettman et al. [17] compared the prevalence of depression symptoms (mild, moderate, moderately severe, severe) according to the PHQ-9 among respondents of the *COVID-19 Life Stressors Impact on Mental Health and Well-being study* conducted in the period March 31, 2020-April 13, 2020 and, like Daly et al. [15], the NHNE Survey conducted in 2017-2018. Results showed that the depression symptom prevalence was higher in every category during COVID-19 than before (mild: 24.6% vs 16.2%, moderate: 14.8% vs 5.7%, moderately severe: 7.9% vs 2.1%, severe: 5.1% vs 0.7%). In addition, in contrast to pre-COVID-19, during COVID-19 moderate to severe depression symptom levels differed between age categories with higher prevalence among younger adults (18-39 years old: 38.8%, 40-59 years old: 26.8%, and ≥60 years old: 14.9%).

## Results prospective studies Netherlands

Three prospective studies in the Netherlands are based on the Dutch *Longitudinal Internet studies for the Social Sciences* (LISS) panel with post-outbreak assessments of mental health in March, May, and June 2020. In the study by Van der Velden et al. [18], like Breslau [16], no increase was found in the prevalence of anxiety and depression symptoms among adults (≥18 years) according to the MHI-5 in March 2020 (17.0%) compared to the pre-outbreak prevalence in November 2019 (16.9%). Compared to 18-34 years old respondents (19.7%), 35-49 years respondents had a significant higher prevalence (22.1%) and 65 years and older respondents had a lower prevalence of symptoms (10.6%) in March 2020. In the period November 2018-March 2019 all three age categories above 34 years old had a lower prevalence than 18-34 years old respondents. The study by Van Tilburg [19] focused on the mental health problems (also according to the MHI-5) among respondents of 65 years and older. Comparisons of separate item scores between November 2019 and May 2020 showed a significant but trivial improvement in mental health (Cohen’s d ≤ 0.18). The study by Van der Velden [20] with data up to June 2020, showed a significant but small decrease in the prevalence of anxiety and depression symptoms (MHI-5) in June (15.3%) compared to March 2020 (17.2%) and November 2019 (16.8%) among adults. In addition, they found that the recovery of symptoms in the period November 2019- March 2020, did not differ significantly from the period March 2020 to June 2020 (remission 16.1% versus 18.0%; improved 5.4% versus 6.0%, unchanged 54.3% versus 56.1%; worsened 24.2% versus 19.9%). This study is part of the ongoing study COVID-19 study using the LISS panel [16, 18, 19].

## Aims current study

In sum, prospective studies in the UK all showed an increase in mental health problems among especially 18-35 years old respondents during the first months after the COVID-19 outbreak compared to previous years. With respect to the US, findings are mixed to some extent. In the prospective study of Breslau et al. [16] among respondents who participated in both waves no differences in mental health problems were found. The other prospective studies [13-17], that extracted data on mental health from different pre- and post-COVID-outbreak samples in the US, showed an increase in the prevalence of mental health problems. Studies using the Dutch population-based LISS panel found no evidence that mental health problems among the Dutch general population increased, although 35-49 years old respondents were more and 65+ respondents were less at risk of mental health problems than 18-34 years old respondents in March 2020.

To what extent the ongoing COVID-19 pandemic and individual and societal consequences affect the mental health of the general population on the longer term (later than June 2020) is unknown. In addition, current prospective studies mainly focused on psychological distress, anxiety, and depression symptoms, and did not provide insight in the effects of the COVID-19 pandemic on the *prevalence* of other relevant mental health problems such as fatigue and sleep problems, and other indicators of mental health such as the use of medicines for anxiety and depression symptoms, use of medicines for sleep problems, use of mental health services, and disabilities due to health problems. Moreover, little is known about the post-COVID-outbreak *incidence* of mental health problems compared to the incidence of mental health problems before the COVID-19 outbreak. Aim of the present prospective population-based study is to fill this gap in scientific knowledge. Current studies suggest that the effects may differ between age categories. We therefore assessed the prevalence and incidence separately among the age categories 18-34 years, 35-49 years, 50-64 years, and 65 years and older. Finally, very little is known about the extent to which prospective risk factors of mental health problems *before* the outbreak are also prospective risk factors of problems and services use after the *outbreak.* Are similar or other groups than “normal” at risk for post COVID-19 outbreak mental health problems and services use? We are not aware of prospective studies addressing this topic, other than Van der Velden et al. [18], that assessed risk factors of anxiety and depression symptoms during the outbreak (March 2020) compared to risk factors of these symptoms one year earlier in March 2019.

For this purpose, data were extracted from surveys in the LISS panel on health conducted in November-December 2018 (T1), November-December 2019 (T2) and November-December 2020 (T3), nine months after the COVID-19 outbreak. Research questions are:

1. To what extent does the *prevalence* of mental health problems and mental health services use among the general population before the COVID-19 outbreak in November- December 2108 (T1) and 2019 (T2), differ from the prevalence after the COVID-19 outbreak in November-December 2020 (T3)?
2. To what extent does the *incidence* of mental health problems and mental health services use among the general population before the COVID-19 outbreak in November-December 2019 (T2), differ from the incidence after the COVID-19 outbreak in November-December 2020 (T3)?
3. To what extent are well-documented *risk factors of* mental health problems and mental health services use before the outbreak (November-December 2019, T2) comparable with risk factors of problems and services use after the outbreak (November-December 2020, T3), such as sex, age, education level, marital status, employment status and physical health [20-23].

Statistics Netherlands (CBS) reported that about 169,000 had died in the Netherlands in 2020, 10% more than expected compared to previous years [26]. This increase is very likely due to COVID-19.

## Materials and methods

### Procedures and participants

For the present study, data on mental health were extracted from the Longitudinal Internet studies for the Social Sciences (LISS) panel [27]. This panel is based on a traditional probability sample drawn from the Dutch population register of 16 years and older by Statistics Netherlands and administered by CentERdata. The set-up was funded by the Dutch Research Council (NWO).

Panel members receive an incentive of 15 euros per hour and members who do not have a computer and/or internet access are provided with the necessary equipment at home (further information about the LISS panel, all conducted studies since 2007, and open access data see: https://www.dataarchive.lissdata.nl; in English). As described above, this panel was also used in earlier mental health-related COVID-19 studies by Van der Velden et al. [18, 20, 21] and Van Tilburg et al. [19].

Data on mental health of adults were extracted from the longitudinal Health module, in particular the waves in November 2018 (T1: N^invited^ =6,466, response=84.4%), in November 2019 (T2: N^invited=^5,954, response=86.4%), and in November 2020 (T3: N^invited^=6,832, response=83.6%). Reminders were sent in December of each year. In total, 4,107 respondents who were 18 years or older at T1, participated at T1, T2, and T3. The total study sample consisted of 4,064 respondents with complete data across the three surveys (99%). We next weighted the data using 16 exclusive demographic profiles among the total adult Dutch population to optimize the representativeness of the current study, based on the data of Statistics Netherlands (see: https://opendata.cbs.nl/#/CBS/en/; in English). The 16 profiles were constructed using the variables sex (male, female), age (18-34, 35-49, 50-64, 65 years and older) and marital status (married and unmarried), yielding 2*4*2=16 demographic profiles. All findings are based on the weighted sample.

### Ethical approval and informed consent

According to the Dutch Medical Research Involving Human Subjects Act (WMO) the present study did not require approval from a Medical Ethical Testing Committee (METC). Nevertheless, the longitudinal Health module (as part of the Longitudinal Core Study in LISS, starting in 2007) were evaluated and approved by the Board of Overseers, an Internal Review Board (IRB) until 2014. In accordance with the General Data Protection Regulation (GDPR), participants gave explicit written consent for the use of the collected data for scientific and policy relevant research. The LISS panel has received the international Data Seal of Approval (see https://www.datasealofapproval.org/ en/). All data of studies conducted with the LISS panel are anonymized and are open access for researchers and policy makers.

### Measures

We used the following six measures administered in each survey to obtain insight in the mental health problems and mental health services use of respondents in November-December 2018, 2019 and 2020. At each survey respondents’ sex, age, marital status and employed status was assessed. For the present study, marital status and employed status were recoded in married (1= yes, 2= no) and employed (1=no, 2= yes).

### Anxiety and depression symptoms

Anxiety and depression symptoms were examined using the Mental Health Index or Inventory (5-item sub scale of the MOS 36-item short-form health survey [28, 29]. Respondents were asked to rate their mental health during the past month on 6-point Likert scales (5=never to 0=continuously). After recoding the positive formulated third and fifth item, the total scores were computed and multiplied by four (all Cronbach’s α> 0.85; range 0 to 100). Lower scores reflect higher symptom levels. A cut-off of≤59 was used to identify respondents with moderate to severe symptom levels and a cut-off of ≤44 to identify respondents with severe symptom levels [30].

### Fatigue and sleep problems

Respondents were furthermore administered a list of 10 (physical) problems people may suffer from, varying from heart complaints to sleeping problems. For the present study we focused on the items ‘*Do you regularly suffer from fatigue’* and ‘*Do you regularly suffer from sleep problems*’ (1=no, 2=yes).

### Disabilities due to (mental) health problems

Psychopathology is consistently and independently associated with increased disability [31]. Health-related disabilities were assessed with the question ‘*To what extent did your physical health or emotional problems hinder your work over the past month, for instance in your job, the housekeeping, or in school?* This question is comparable with questions of the MOS-36 [28] and the European Health Interview Surveys (SILC-EU) [32], and had 5-point Likert scale (1=not at all to 5=very much). For the present study scores were recoded into low (1=1,2,3) and high (2=4,5).

### Medicines for anxiety and depression

Use of medicines were assessed for various conditions varying from medicines for blood pressure to sleep problems ( ‘*Are you currently taking medicine at least once a week for* .. ’). For the present study we focused on current use of medicines for anxiety and depression symptoms and sleep problems (0=no, 1=yes).

### Mental health services use

The use of mental health services (MHS) was assessed by one question ‘*How often did you use the following health services over the past 12 months*?’, with answer categories varying from psychiatrist/ psychologist/ psychotherapist to dentist. For the present study, we focused on the use of a psychiatrist, psychologist, or psychotherapist and recoded ‘use’ into ‘no’ (0=no use) and ‘yes’ (1=once or more) in the past 12 months.

### Physical disease

Respondents were administered a list of 19 physical diseases/ problems and asked if they had one of more of these diseases/problems according to a physician (‘*Has a physician told you this last year that you suffer from one of the following diseases/ problems?*’) varying from cancer to a chronic lung disease. For the represent study we recoded the answers into ‘no disease’ (1=none of the diseases/problems included in ‘diseases’) and ‘disease’ (including: (2= angina, pain in the chest a heart attack including infarction or coronary thrombosis or another heart problem including heart failure; a stroke or brain infarction or a disease affecting the blood vessels in the brain diabetes or a too high blood sugar level; chronic lung disease such as chronic bronchitis or emphysema; asthma; arthritis, including osteoarthritis, or rheumatism, bone decalcification or osteoporosis; cancer or malignant tumor, including leukemia or lymphoma; and/or benign tumor (skin tumor, polyps, angioma)).

### Data analyses

To examine the extent to which the prevalence of the seven assessed mental health problems after the COVID-19 outbreak in November-December 2020 (T3) changed compared to the prevalence in November-December 2018 (T1) and 2019 (T2), repeated measures multivariate logistic regression (RMMLR) analyses (GEE) were conducted with problems at T1, T2 and T3 as dependent variables (separate analyses for each dependent variable). In the analyses sex, age, marital status, employment status, education level and disease at T1, T2 and T3 were used as control variables.

For the incidence at T3 (prevalence of new cases), the prevalence of mental health problems and use at T3 was assessed among those *without* these problems and or use at T2. Likewise, for the incidence at T2, problems and use at T2 were assessed among those *without* these problems and use at T1 (the incidence at T1 could not be computed among the current study sample). To assess the extent to which the incidence of mental health problems and use in November-December 2020 (T3) changed compared to the incidence in November-December 2019 (T2), multivariate logistic regression were conducted as follows. The incidence at T2 and T3 cannot directly be compared because those with problems and use at T2 and T3 without problems and use at the previous survey partly overlap. To enable a comparison of the incidence at T2 and T3, we therefore first randomly split the total study sample into two almost equal independent subgroups of respondents (A: n=2,025; and B: n=2,019; because of the weighting the numbers of both groups slightly differs). Of both subgroups the incidence at T2 (A1, B1) and T3 (A2, B2) were computed. We finally compared the incidence of A1 with B2 and compared the incidence of B1 with A2 using multivariate logistic regression with the same control variables as in the RMMLR analyses (on T1 for analyses incidence at T2, on T2 for analyses incidence at T3).

Prospective risk factors of mental health problems and services use were examined using multivariate logistic regression analyses with problems and services use at T2 and T3 as dependent variables. The variables sex, age, education level, marital status, employment status and physical disease at T1 and T2 respectively were simultaneously entered as predictors (for instance, variables at T2 were entered as predictors for problems at T3).

## Results

### Non-response

Multivariate logistic regression analyses (before weighting) with non-response at T2 and T3 as dependent variable (1= participated at T1, T2 and T3, 2=did not participate at T2 and T3) showed that the non-response was not significantly (p <0.05) associated with the seven mental health and mental services use variables at T1, disease at T1, and not with sex, employment status, and education level at T1. Unmarried respondents (85.1%) compared to married respondents (90.4%) participated less at T1, T2 and T3 (adjusted Odds ratio (aOR)=0.80, 95% confidence interval (95% CI)=0.66-0.98, p=0.029)). Compared to 18-34 years old respondents (79.1%), 34-50 years old (85.7%, aOR=1.43, 95% CI=1.10-1.87, p=0.008), 50-64 years old (92.3%, aOR=2.80, 95% CI=2.10-3.75; p<0.001), and 65 years or older respondents (90.7%, aOR=2.10, 95% CI=1.53- 2.84, p<.001) participated significantly more often. As described, the study sample was weighted for, besides employment status, marital status and age.

### Characteristics respondents

The characteristic of the study sample are presented in Table 1. It shows, among others, that in absolute numbers as time passes by more respondents are married, become older, have a higher education level, as well as more often have a physical disease. At T3 the youngest respondents were 20 years old.

**Table 1.**
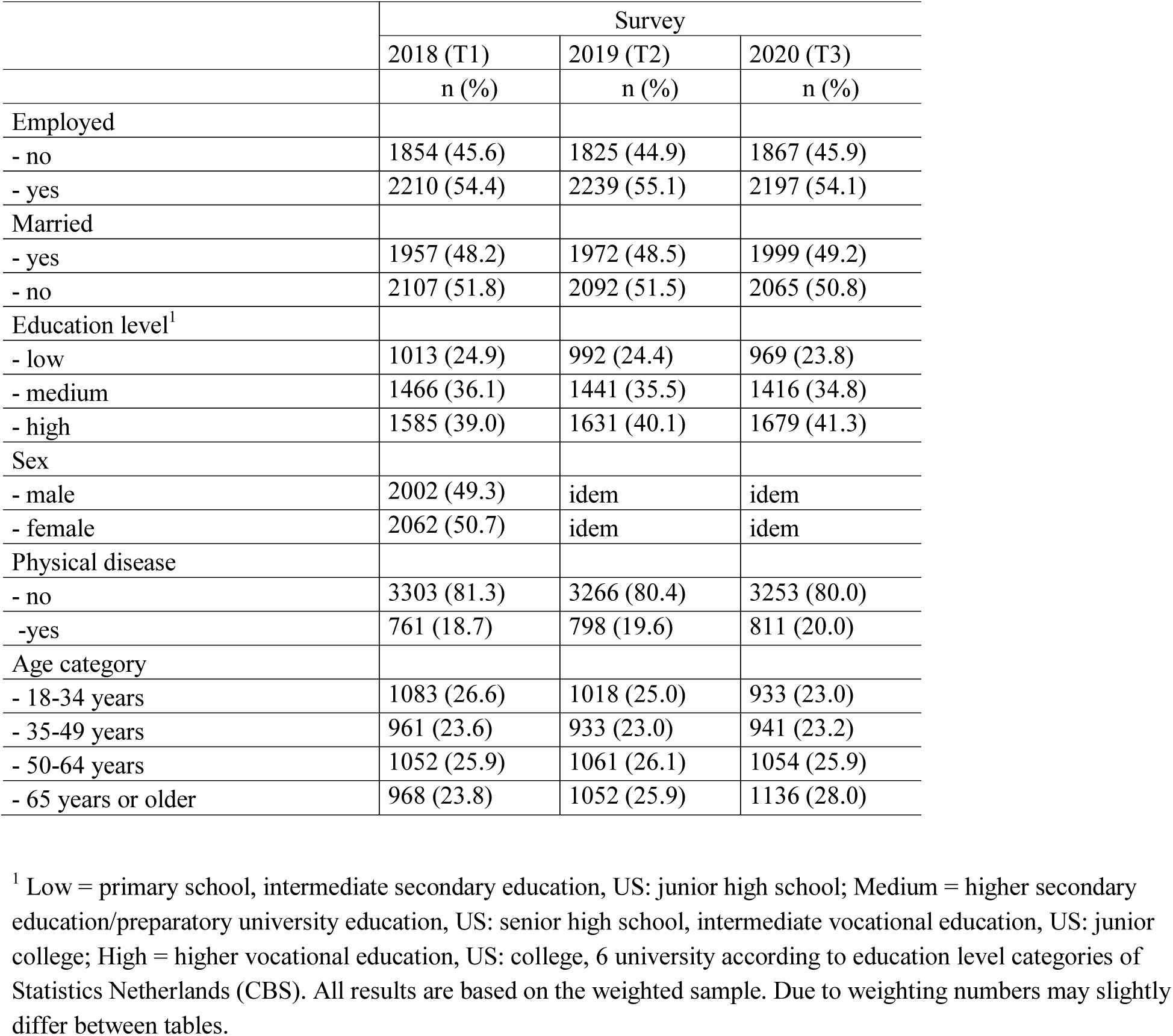
Characteristics respondents (N=4,064)

|  | Survey |  |  |
| --- | --- | --- | --- |
|  | 2018 (T1) | 2019 (T2) | 2020 (T3) |
|  | n (%) | n (%) | n (%) |
| Employed |  |  |  |
| - no | 1854 (45.6) | 1825 (44.9) | 1867 (45.9) |
| - yes | 2210 (54.4) | 2239 (55.1) | 2197 (54.1) |
| Married |  |  |  |
| - yes | 1957 (48.2) | 1972 (48.5) | 1999 (49.2) |
| - no | 2107 (51.8) | 2092 (51.5) | 2065 (50.8) |
| Education level<sup>1</sup> |  |  |  |
| - low | 1013 (24.9) | 992 (24.4) | 969 (23.8) |
| - medium | 1466 (36.1) | 1441 (35.5) | 1416 (34.8) |
| - high | 1585 (39.0) | 1631 (40.1) | 1679 (41.3) |
| Sex |  |  |  |
| - male | 2002 (49.3) | idem | idem |
| - female | 2062 (50.7) | idem | idem |
| Physical disease |  |  |  |
| - no | 3303 (81.3) | 3266 (80.4) | 3253 (80.0) |
| -yes | 761 (18.7) | 798 (19.6) | 811 (20.0) |
| Age category |  |  |  |
| - 18-34 years | 1083 (26.6) | 1018 (25.0) | 933 (23.0) |
| - 35-49 years | 961 (23.6) | 933 (23.0) | 941 (23.2) |
| - 50-64 years | 1052 (25.9) | 1061 (26.1) | 1054 (25.9) |
| - 65 years or older | 968 (23.8) | 1052 (25.9) | 1136 (28.0) |

<sup>1</sup> Low = primary school, intermediate secondary education, US: junior high school; Medium = higher secondary education/preparatory university education, US: senior high school, intermediate vocational education, US: junior college; High = higher vocational education, US: college, 6 university according to education level categories of Statistics Netherlands (CBS). All results are based on the weighted sample. Due to weighting numbers may slightly differ between tables.

### Prevalence mental health problems total study sample

The prevalence of the assessed mental health problems and services use among the total study sample at T1, T2 and T3 are presented in Table 2. According to the repeated measures multivariate logistic regression analyses, the prevalence of mental health problems and services use at T3 did not significantly differ from T1 and T2 except for disabilities due to health problems. The prevalence of disabilities due to health problems was significantly lower at T3 (8.5%) than at T1 (9.9%), but not significantly different from the prevalence at T2 (8.5%). Analyses for anxiety and depression scores using the cut-off score of ≤44 of the MHI-5 showed similar outcomes: the prevalence of severe symptom levels at T1 (6.6%), T2 (6.5%) and T3 (6.3%) did not differ significantly.

**Table 2.** Prevalence of mental health problems and services use (N=4,064)

|  | November-December 2018 (T1) | November-December 2019 (T2) | November-December 2020 (T3) | T1 versus T3 | T2 versus T3 | T1 versus T2 |
| --- | --- | --- | --- | --- | --- | --- |
| **Prevalence** | n (%) | n (%) | n (%) | aOR (95% CI) | aOR (95% CI) | aOR (95% CI) |
| Anxiety and depression symptoms<sup>1</sup> | 660 (16.2) | 687 (16.9) | 686 (16.9) | 1.09 (0.99-1.19) | 1.00 (0.92-1.09) | 1.08 (0.99-1.18) |
| Sleep problems | 834 (20.5) | 871 (21.4) | 869 (21.4) | 1.03 (0.98-1.09) | 0.99 (0.95-1.03) | 1.05 (1.00-1.09)* |
| Fatigue | 1275 (31.3) | 1292 (31.8) | 1266 (31.2) | 1.00 (0.96-1.05) | 0.98 (0.95-1.01) | 1.02 (0.99-1.06) |
| Disabilities due to health problems | 404 (9.9) | 345 (8.5) | 348 (8.6) | 0.86 (0.76-0.97)* | 1.03 (0.90-1.17) | 0.84 (0.74-0.95)** |
| Medicines for anxiety/depression | 190 (4.7) | 187 (4.6) | 195 (4.8) | 1.04 (0.96-1.13) | 1.04 (0.98-1.11) | 1.00 (0.94-1.06) |
| Medicines for sleep problems | 189 (4.6) | 203 (5.0) | 207 (5.1) | 1.05 (0.94-1.17) | 1.00 (0.91-1.09) | 1.05 (0.96-1.15) |
| Use of mental health services<sup>2</sup> | 341 (8.4) | 345 (8.5) | 317 (7.8) | 0.95 (0.83-1.09) | 0.92 (0.82-1.04) | 1.03 (0.91-1.16) |

<sup>1</sup>According cut-off score of $\leq 59$ on MHI-5. <sup>2</sup>Psychiatrist/psychologist/psychotherapist in past 12 months. aOR=Odds ratio of repeated measures logistic regression analyses, adjusted for sex, age, marital status, employment status, education level and disease at T1, T2, and T3. 95% CI=95% confidence interval for aOR. All results are based on the weighted sample. Due to weighting numbers may slightly differ between tables.
\* $p < 0.05$ , \*\* $p < 0.01$

### Prevalence mental health problems and services use among age categories

Among respondents 18-34 years old, the prevalence of anxiety symptoms at T3 (22.3%) was significantly higher than at T1 ((19.9%, adjusted Odds Ratio (aOR)=1.30, 95% Confidence interval (95% CI)=1.08-1.57, p=0.009)), but not compared to T2 (20.7%). In contrast, among respondents 50-64 years old the prevalence of these symptoms at T3 (13.3%) was significantly lower than at T1 (14.6%, aOR=0.82, 95% CI=0.68-0.99, p=0.043) and T2 (15.1%; aOR=0.81, 95% CI=0.68-0.95, p=0.013).

With respect to sleep problems, among respondents 18-34 years old the prevalence at T3 (14.6%) was significantly higher than at T1 (13.2%; aOR =1.19, 95% CI=1.02-1.39, p=0.023), but not compared to T2 (14.8%).

For fatigue, no significant changes in the prevalence were found between T1, T2 and T3 across the four age categories. Only among respondents 50-64 years old the prevalence of disabilities due to health problems differed significantly to some extent: the prevalence was lower at T3 (10.6%) compared to T1 (12.2; aOR=0.78, 95% CI= 0.62-0.99, p=0.039, but not compared to T2 (10.0%).

The use of medicine for anxiety and depression among respondents 18-34 years old was significant more prevalent at T3 (3.4%) than at T2 (2.9%; aOR=1.21, 95% CI=1.01-1.46, p=0.040) but not at T1. Concerning the use of medicines for sleep problems, no significant changes in the prevalence were found between T1, T2 and T3 across the four age categories.

Finally, the use of mental health professionals among respondents of 65 years old and older decreased significantly in the 12 months before T3 (2.2%) compared to T1 (3.5%; aOR=0.60, 95% CI=0.41-0.87, p=0.006) and T2 (3.1%; aOR=0.69, 95% CI=0.49-0.97, p=0.031). No significant differences were found between T1 and T2 (for details, see S1 Appendix Age categories).

### Incidence mental health problems total study sample

Table 3 shows the incidence of mental health problems and services use at T2 and T3 among the total study sample and the results of crosswise multivariate logistic regression analyses, showing that the incidence of the mental health problems and services use at T2 and T3 did not differ significantly. Similar analyses using the cut-off ≤44 of the MHI-5 showed that the incidence of severe anxiety and depression symptom levels at T2 (3.7%) and T3 (3.3%) did not significantly differ (results not shown in table).

**Table 3.** Incidence of mental health problems (N=4,064)

|  | November-December<br>2018 (T1) | November-December<br>2019 (T2) | November-December<br>2020 (T3) | A1 versus B2 | B1 versus A2 |
| --- | --- | --- | --- | --- | --- |
| **Incidence<sup>1</sup>** | n (%) | n (%) | n (%) | aOR (95% CI) | aOR (95% CI) |
| Anxiety and depression symptoms<sup>2</sup> | n.a. | 293 (8.6) | 274 (8.1) | 1.00 (0.78-1.29) | 0.95 (0.75-1.22) |
| Sleep problems | n.a. | 120 (3.7) | 93 (2.9) | 0.86 (0.59-1.25) | 0.71 (0.47-1.07) |
| Fatigue | n.a. | 112 (4.0) | 93 (3.4) | 0.79 (0.53-1.19) | 0.87 (0.59-1.28) |
| Disabilities due to health problems | n.a. | 173 (4.7) | 205 (5.5) | 1.34 (1.00-1.80) | 1.06 (0.78-1.42) |
| Medicines for anxiety/depression | n.a. | 14 (0.4) | 20 (0.5) | 1.63 (0.56-4.77) | 1.31 (0.54-3.20) |
| Medicines for sleep problems | n.a. | 38 (1.0) | 36 (0.9) | 1.19 (0.64-2.21) | 0.76 (0.38-1.54) |
| Use of mental health services | n.a. | 158 (4.2) | 136 (3.7) | 0.99 (0.72-1.38) | 0.77 (0.55-1.09) |

<sup>1</sup>Prevalence mental health problems among those without problems /use on previous survey.
<sup>2</sup>According to cut-off score of $\leq 59$ on MHI-5.
<sup>3</sup>Psychiatrist/psychologist/psychotherapist in past 12 months. n.a.=not available because 2017 survey was outside aim of this study. aOR=Odds ratio of logistic regression analyses, adjusted for sex, age, marital status, employment status, education level and disease at T1 and T2, and T2 and T3. 95% CI= 95% confidence interval for aOR. A1=incidence T2 of subgroup A. B1=incidence T2 of subgroup B. A2=incidence T3 of subgroup A. B2=incidence T3 of subgroup B. All results are based on the weighted sample. Due to weighting numbers may slightly differ between tables.

### Incidence mental health problems among age categories

Due to the cell counts of the incidence of mental health problems and services use at T2 and T3 (see Table 3) we limited the (cross-wise) analyses multivariate logistic regression analyses among the four age categories to the incidence of anxiety and depression symptoms. The events-per-variable (EPV) ratio for the other dependent variables, with seven predictors including control variables, became lower than 10. The results of these analyses showed no significant differences in incidence between T2 and T3 among 18-34 years old respondents (T2^incidence^=12.4%, T3^incidence^=13.1%), 35-49 years old respondents (T2^incidence^=9.9%, T3^incidence^ = 9.7%), 50-64 years old respondents (T2^incidence^=6.2%, T3^incidence^=5.0%), and 65 years and older respondents (T2^incidence^=6.1%, T3^incidence^=4.8%). Using the cut-off ≤ 44 of the MHI-5 also did not reveal significant differences between the T2 and T3 incidence (results not shown in table).

### Risk factors of mental health problems and services use

In Table 4, the results of the multivariate logistic regression analyses are presented (for 95% confidence intervals of the adjusted OR’s, see S2 Appendix Risk factors). Table 4 shows that existing mental health problems and services use (e.g., assessed one year earlier) were by far the strongest predictors for problems and use before and after the COVID-19 outbreak. For example, about 90% of the respondents with existing sleep problems and fatigue T1 and T2, had sleep problems and fatigue at T2 and T3, respectively. With respect to anxiety and depression symptoms at T2 and T3, almost the same predictors were significant for these symptoms at T2 and T3: only those with a medium education level were no longer less at risk of symptoms than those with a relatively low education level. An almost similar pattern can be observed for sleep problems: only males and females did no longer differ in sleep problems after the outbreak (T3). However, 35-49 years old respondents were more at risk of sleep problems after the outbreak (at T3), but not before (at T2), relative to 65+ respondents. The patterns of risk factors of fatigue at T2 and T3 are identical. For disabilities due to health problems, e.g. that physical health or emotional problems hinder respondents work over the past month, for instance in their job, housekeeping, or in school, results show that significant differences between subgroups disappeared after the outbreak: those with a medium and high education level compared to those with a relative low level, males compared to females, and youngest adult group (18-34 years) compared to 65+ respondents no longer differed in disabilities due to health problems. Finally, for mental health services use those employed before the outbreak (T2) used services significantly less often services after the outbreak (T3) than unemployed, in contrast to the period before the outbreak (T1-T2). Because of the relative low prevalence of medicines for anxiety/depression and medicines for sleep problems, we have omitted these dependent variables from the analyses.

**Table 4.**
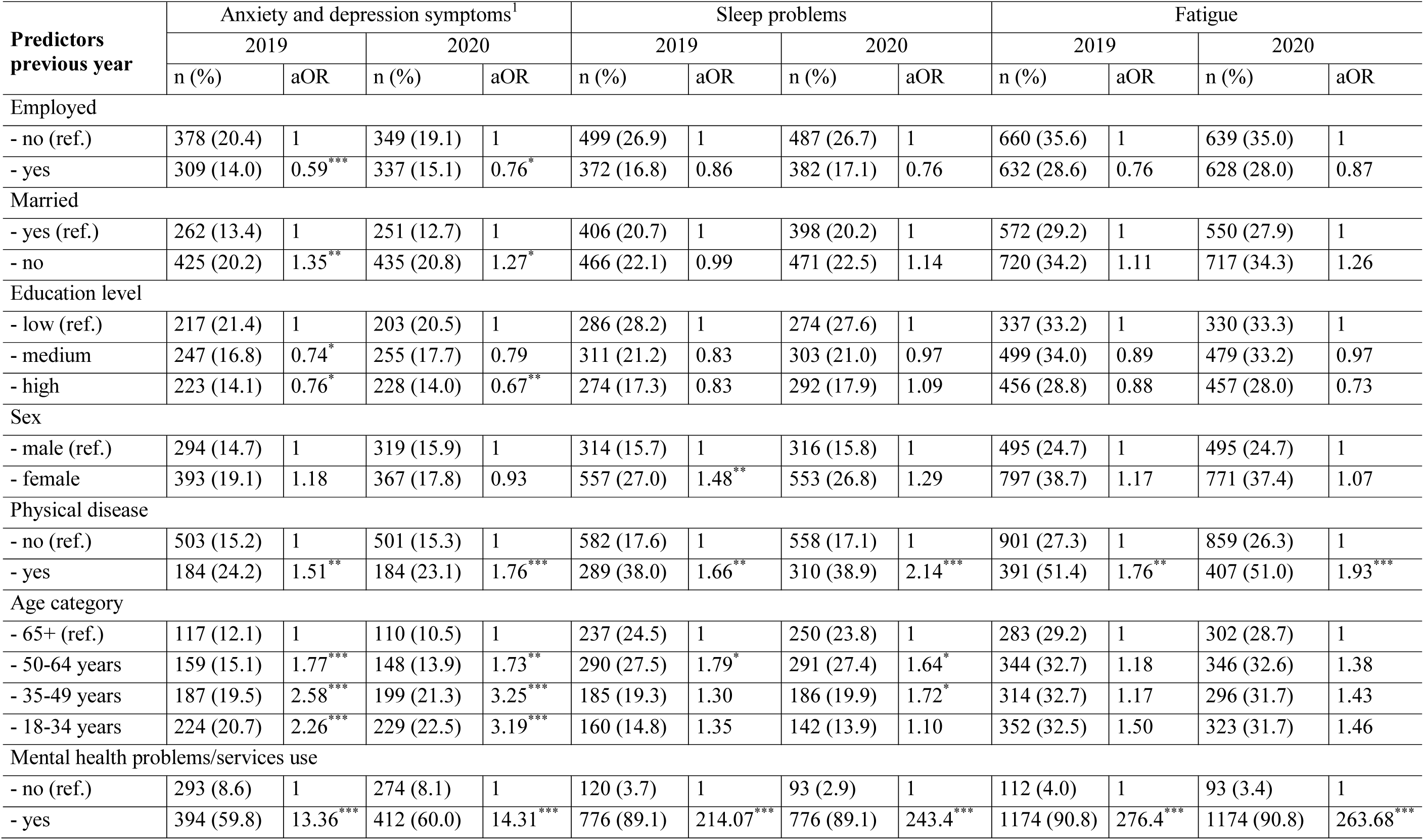
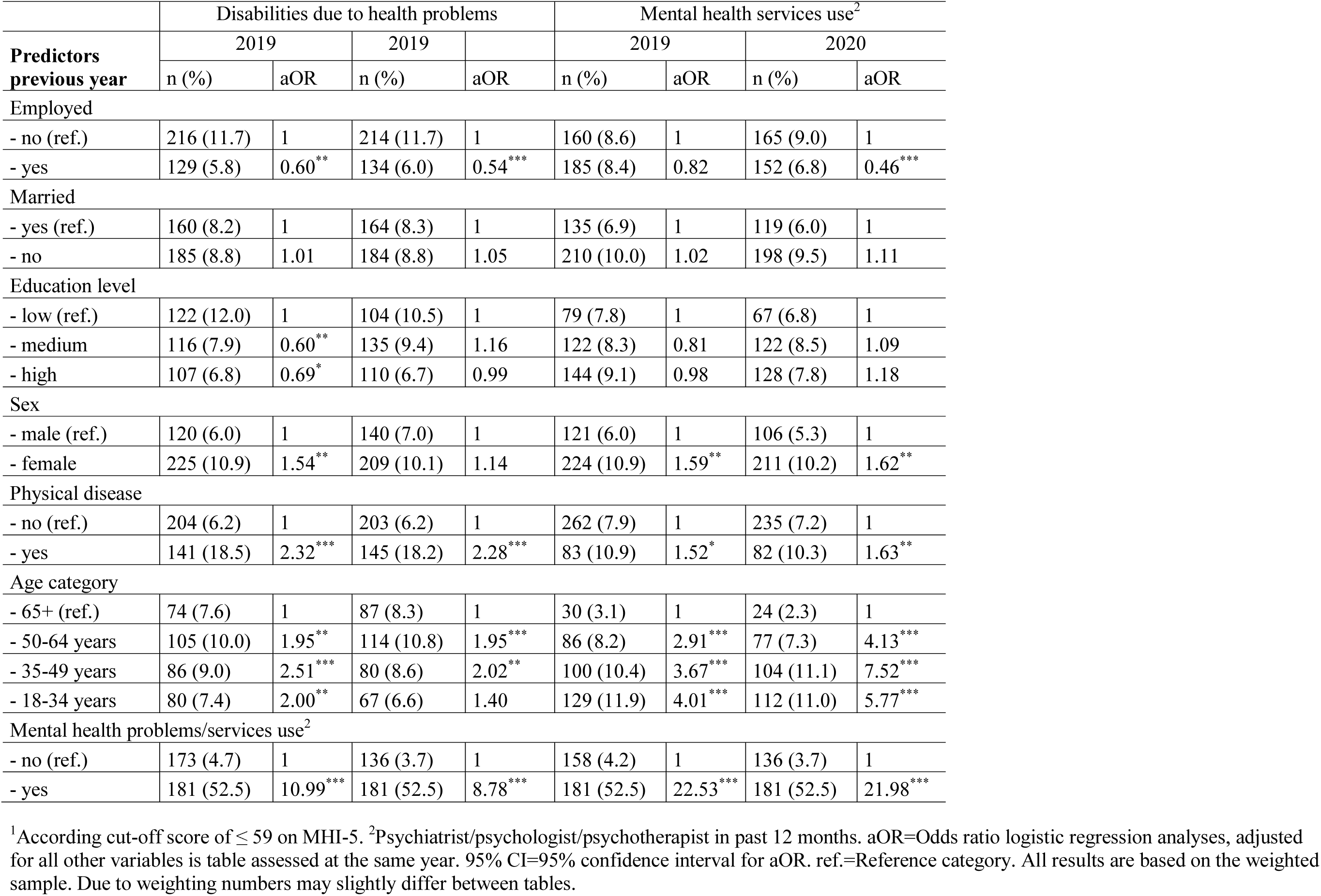
Risk factors of mental health problems and services use (N=4,064)

## Discussion

The first aim of the present COVID-19 study was to examine the *prevalence* of mental health problems and mental health services use among the adult general population 9 months after the COVID-19 outbreak (November-December 2020, T3), compared to the prevalence in similar periods before the outbreak (November-December 2018 and 2019, T1 and T2). The second aim was to examine the *incidence* of mental health problems and mental health services use between November-December 2019 and November-December 2020, compared to the incidence between November-December 2018 and November-December 2019. The third aim was to examine *risk factors of* mental health problems and services use 9 months after the outbreak, compared to risk factors of mental health problems and services use before the outbreak (November-December 2019). For these purposes, data were extracted from the LISS panel that is based on a large probability sample of the Dutch population. To optimize the representativeness of the study sample (N=4,064), data were weighted using 16 demographic profiles of the adult Dutch population.

The main conclusion that can be drawn from the present study is that the prevalence, the incidence as well as risk factors of the assessed mental health problems and services use appear to be very stable among the general population (cf. [33]). We found no indications that on population level the prevalence nor the the incidence of anxiety and depression symptoms, sleep problems, fatigue, disabilities due to health problems, use of medicines for sleep problems, anxiety and depression, and mental health services use increased in November-December 2020, compared to pre-COVID-19 mental health and services use in November-December 2018 and in 2019. Based on the results of previous COVID-19 studies, we also examined the prevalence among the 18-34 years, 35-49 years, 50-64 years, and 65 years and older respondents separately, showing almost identical results. Results did not show differences between mental health problems and mental health services use at T1 and T2 on the one side, and problems and use at T3 on the other side, except among 50-64 years old respondents and among 65+ old respondents. Results showed a small significant *decrease* in the prevalence of anxiety and depression symptoms at T3 (13.3%) compared to T1 (14.6%) and T2 (15.1%), and a small significant *decrease i*n the use of mental health services at T3 (2.2%) compare to T1 (3.5%) and T2 (3.1%). Our results about mental health problems seem to differ from almost all identified COVID-19 studies in the UK and US, but these studies focused on mental health problems until the summer of 2020.

Importantly, in our previous study [20] assessing anxiety and depression symptoms until the summer of 2020 (June), like the study of Breslau et al. [16] and Hyland et al. [34], no increase in prevalence compared to November-December 2019 was observed. Van Tilburg reported a similar finding et al. [19] among older respondents. As described, in the study of Breslau et al. [16] also no differences between the past-month prevalence of serious psychological distress in May 2020 and past-year prevalence assessed in February 2019 were found. In addition, they found an incidence of 13.7% when distinguishing K6 scores in low-distress versus mild/moderate-serious distress, and an incidence of 4.9% when distinguishing K6 scores in low- mild/moderate distress versus serious distress. The last incidence number, although we used the MHI-5, seem to be close to our incidence number for severe anxiety and depression symptom levels at T2 (3.7%) and T3 (3.3%). The extent to which the incidence of mental health problems after the COVID-19 outbreak in the UK and US differed from the incidence before the outbreak is unknown. Nevertheless, in our previous study [18] we found no indications that the recovery of symptoms after the outbreak differed from the recovery before the outbreak. We are not aware of COVID-19 studies to compare our findings on sleep problems, fatigue, use of pharmaceuticals, and use of mental health services use with.

How can we, given the outcomes of several studies showing an increase in mental health problems [9-17], understand the contradicting results of the present study? In addition, in our previous study [18] we observed a significant increase in emotional loneliness which is considered a risk factor for future mental health problems [33, 34]. Current findings indicate that, despite the observed increase in loneliness until the summer of 2020, the prevalence of anxiety and depression symptoms did not increase among the general population nor in different age categories.

Several elements, other than the resilience of people [3-5], may play a role and may help explain these differences, but future empirical research is required to examine and test these elements and explanations. First, unemployment might be a cause for increases in mental health problems [35] but unemployment remained relatively stable in the Netherlands during the COVID-19 outbreak whereas they increased strongly in the US in March and April 2020. However, also in the UK the unemployment figures did not rise as dramatically as in the US in the first two months after the outbreak. From May 2020 on, due to the COVID-19 recovery programs put in place, the unemployment figures declined again strongly in the US whereas they remained stable in the UK and the Netherlands [38]. Hence, the unemployment incidence might explain some of the differences with the US results but they cannot explain the differences with the UK findings, even though it might still be that support in the UK for mental health problems is poorer than in the Netherlands. A recent report by Statistics Netherlands [39] showed that in the period July 2020-September 2020, 83% of the Dutch resident were positive about their general health, compared to 79-80% in the same period in 2017, 2018 and 2019. In addition, the study by De Vries et al. [40] among members of the Dutch *Health Care Consumer Panel* between March 16 and May 17 2020, concluded that members were positive about the mitigation measures, that they trusted the information and the measures from authorities, and adopted protective measures.

However, as said Breslau et al. using pre- and post-outbreak data of the same US study sample, did not found an increase of mental health problems compared to the worst months before the outbreak [16]. Analyses of the UK Household Longitudinal Study data, showed an increase until the summer of 2020 in contrast to the Netherlands. Another possible explanation could be that the Brexit process and final end of the UK membership of the EU in 2020, similar to the presidential elections, strongly divided the country resulting in societal tensions and uncertainties about the future outside the EU. These tensions and uncertainties may have increased the risk of mental health problems. With respect to the political climate, other than the US where the COVID-19 pandemic became part of an intense political debate and elections, to date this pandemic did not result in a similar political discourse in the Netherlands as in the US.

Furthermore, mental disorders in the US were much more prevalent in the US than in the Netherlands which may increase the risk for higher post-outbreak mental health problems [41]. It might be that the Dutch welfare system provide better support to people with mental health issues during the COVID-19 outbreak or to people who ran into mental health issues after the outbreak than the US or UK welfare system does.

### The role of pre-COVID mental health and stressors

The absence of an increase of mental health problems between T1 and T2 on the one side and T3 on the other side does not indicate that the COVID-19 outbreak has not negatively affected the mental health of individuals. As shown by the incidence rates, a minority suffer from mental health problems not experienced one year earlier that may be partly related to the disruptive effects of the COVID-19 pandemic. In addition, besides this pandemic, on a yearly basis about 40% of the adults is confronted with potentially traumatic and life-events that increase the risk for mental health problems among those confronted with these events [42, 43]. There are no valid reasons to assume that those events put the mental health less at risk because of this COVID-19 pandemic. The stress sensitization model suggests otherwise [44]. In other words, without ignoring the drastic societal effects of this pandemic we should be aware this pandemic does not occur in a vacuum. The very strong predictive values of assessed previous mental health problems clearly demonstrate, as in the study by Breslau et al. [16], this aspect. Perhaps this pandemic partly reveals mental health problems and mental health problems patterns that were already present before the pandemic but, because of the large (media and scientific) attention towards the mental health effects [7, 45], become more visible because of the pandemic. For example, the number of studies indexed in Pubmed that can be retrieved with the keyword depression is 28,762 with publication date 2018, 30,232 with publication date 2019, and 35,571 with publication date. In 2020, 2,073 were corona or COVID-19 related. An increased visibility of mental health problems should not be confused with an (strong) increased prevalence of mental health problems. In either way, this underscores the relevance of non-retrospective pre-COVID mental health assessment among post-COVID-19 population-based probability samples.

### Strengths and limitations

Although our results showed very clear patterns, some limitations need to be discussed when interpreting and using the outcomes of this study. We did not conduct clinical interviews that would certainly have enriched our study. Although our results do not point in the direction of a strong post-COVID-19 outbreak increase of mental disorders, future studies using clinical interviews will provide further insight in this topic. We extended previous COVID-19 studies by assessing sleep problems, fatigue, use of medicines for sleep problems, anxiety and depression, disabilities due to health problems, and mental health services utilization. However, future studies focusing on other mental health problems such as eating problems, panic attacks, phobias, alcohol and drug misuse, and low self-esteem are warranted. The three surveys had a one-year time interval. Although we examined anxiety and depression symptoms in March 2019, March 2020 and June 2020 in previous studies, we cannot rule out the possibility that significant increases and decreases can be observed using shorter time intervals. The last survey in the present study was conducted in November-December 2020. How this pandemic will develop in the next year is uncertain. Monitoring of mental health of the general population is therefore needed, also because the duration of this pandemic on the longer term may undermine the capacities of individuals to cope with the consequences. It is unclear to what extent the results can be generalized to other developed countries who were hit harder by the COVID-19 pandemic. Finally, this study focused on adults and did not include children and adolescents who might be affected differently. At T3, the youngest respondent was 20 years old. Nevertheless, the prospective study design, the use of a large population-based probability sample, the high response rates, a non-response nor being related to our dependent variables, and the weighting of data using 16 demographic profiles of the Dutch population are major strengths of the present study.

## Data Availability

The study was conducted using the Dutch Longitudinal Internet studies for the Social Sciences (LISS) panel. Further information about all conducted surveys and regulations for free access to the data can be found at https://www.dataarchive.lissdata.nl/ (in English). The LISS panel has received the international Data Seal of Approval (see https://www.datasealofapproval.org/en/).
All data of studies conducted with the LISS panel are anonymized. For this study we used the open access data of the Health module (see https://www.dataarchive.lissdata.nl/study_units/view/12, wave 11, 12 and 13).

## Notes

### Competing Interest Statement

The authors have declared no competing interest.

### Clinical Trial

not applicable

### Funding Statement

No external funding received

### Author Declarations

IRB. According to the Dutch Medical Research Involving Human Subjects Act (WMO) the present study did not require approval from a Medical Ethical Testing Committee (METC). Nevertheless, the longitudinal Health module (as part of the Longitudinal Core Study in LISS, starting in 2007) were evaluated and approved by the Board of Overseers, an Internal Review Board (IRB) until 2014. In accordance with the General Data Protection Regulation (GDPR), participants gave explicit written consent for the use of the collected data for scientific and policy relevant research. The LISS panel has received the international Data Seal of Approval (see https://www.datasealofapproval.org/en/). All data of studies conducted with the LISS panel are anonymized. OPEN ACCESS. The data of the LISS panel are open access for researchers and policy makers. Further information about the regulations for free access to the data can be found at https://www.dataarchive.lissdata.nl. Access to the data was granted after registration.

## References

1. WHO. Coronavirus disease (COVID-19) Weekly Epidemiological Update and Weekly Operational Update. https://www.who.int/emergencies/diseases/novel-coronavirus-2019/situation-reports (accessed February 10, 2021)

2. WHO. Coronavirus disease (COVID-19): Vaccines. https://www.who.int/news-room/q-a-detail/coronavirus-disease-(covid-19)-vaccines (accessed February 10, 2021).

3. Hobfoll SE. Conservation of resources. A new attempt at conceptualizing stress. Am Psychol. 1989; 44: 513-24. https://doi.org/10.1037//0003-066x.44.3.513

4. Hobfoll SE. Social and psychological resources and adaptation. Rev Gen Psychol. 2002; 6: 307-324. https://doi.org/10.1037//1089-268064307

5. Chen S, Bonanno GA. Psychological adjustment during the global outbreak of COVID-19: A resilience perspective. Psychol Trauma 2020; 2: s51–s54. https://doi.org/10.1037/tra000068510.1037/tra0000685

6. Wessely S. Don’t panic! Short and long term psychological reactions to the new terrorism: the role of information and the authorities. J Mental Health 2005; 14: 1-6. https://doi.org/10.1080/09638230500048099

7. Prati G, Mancini AD. The psychological impact of COVID-19 pandemic lockdowns: a review and meta-analysis of longitudinal studies and natural experiments. Psychol Med. 2021; 1-11. https://doi.org/10.1017/S0033291721000015

8. Santabárbara J, Lasheras I, Lipnicki DM, Bueno-Notivol J, Pérez-Moreno M, López-Antón R, De la Cámara C, Lobo A, Gracia-García P. Prevalence of anxiety in the COVID-19 pandemic: An updated meta-analysis of community-based studies. Prog Neuropsychopharmacol Biol Psychiatry. 2020; 109:110207. https://doi.org/10.1016/j.pnpbp.2020.110207

9. Pierce M, Hope H, Ford T, Hatch S, Hotopf M, John A, Kontopantelis E, Webb R, Wessely S, McManus S, Abel KM. Mental health before and during the COVID-19 pandemic: a longitudinal probability sample survey of the UK population. Lancet Psychiatry. 2020; 7: 883-892. https://doi.org/10.1016/S2215-0366(20)30308-4

10. Proto E, Quintana-Domeque C. COVID-19 and mental health deterioration by ethnicity and gender in the UK. PLoS One. 2021; 16(1): e0244419. https://doi.org/10.1371/journal.pone.0244419

11. Niedzwiedz CL, Green MJ, Benzeval M, Campbell D, Craig P, Demou E, Leyland A, Pearce A, Thomson R, Whitley E, Katikireddi SV. Mental health and health behaviours before and during the initial phase of the COVID-19 lockdown: longitudinal analyses of the UK Household Longitudinal Study. J Epidemiol Community Health. 2020; 25: jech- 2020-215060. https://doi.org/10.1136/jech-2020-215060

12. Daly M, Sutin AR, Robinson E. Longitudinal changes in mental health and the COVID- 19 pandemic: evidence from the UK Household Longitudinal Study. Psychol Med. 2020; 13:1-10. https://doi.org/10.1017/S0033291720004432

13. Twenge JM, Joiner TE. U.S. Census Bureau-assessed prevalence of anxiety and depressive symptoms in 2019 and during the 2020 COVID-19 pandemic. Depress Anxiety 2020; 1–3. https://doi.org/10.1002/da.23077

14. McGinty EE, Presskreischer R, Han H, Barry CL. Psychological distress and loneliness reported by US adults in 2018 and April 2020. JAMA. 2020; 324: 93-94. https://doi.org/10.1001/jama.2020.9740

15. Daly M, Sutin A, Robinson E. Depression reported by US adults in 2017–2018 and March and April 2020. J Affect Disord. 2020; 278: 131-135. https://doi.org/10.1016/j.jad.2020.09.065

16. Breslau J, Finucane ML, Locker AR, Baird MD, Roth EA, Collins RL. A longitudinal study of psychological distress in the United States before and during the COVID-19 pandemic. Prev Med. 2021; 43: 106362. https://doi.org/10.1016/j.ypmed.2020.106362

17. Ettman CK, Abdalla SM, Cohen GH, Sampson L, Vivier PM, Galea S. Prevalence of Depression Symptoms in US Adults Before and During the COVID-19 Pandemic. JAMA Netw Open. 2020; 3(9): e2019686. https://doi.org/10.1001/jamanetworkopen.2020.19686

18. van der Velden PG, Contino C, Das M, van Loon P, Bosmans MWG. Anxiety and depression symptoms, and lack of emotional support among the general population before and during the COVID-19 pandemic. A prospective national study on prevalence and risk factors. J Affective Disord. 2020; 277: 540-548. https://doi.org/10.1016/j.jad.2020.08.026

19. van Tilburg TG, Steinmetz S, Stolte E, van der Roest H, de Vries DH. Loneliness and mental health during the COVID-19 pandemic: A study among Dutch older adults. J Gerontol B Psychol Sci Soc Sci. 2020; gbaa111. https://doi.org/10.1093/geronb/gbaa111

20. van der Velden PG, Hyland P, Contino C, von Gaudecker HM, Muffels R, Das M. Anxiety and depression symptoms, the recovery from symptoms, and loneliness before and after the COVID-19 outbreak among the general population: Findings from a Dutch population-based longitudinal study. PLoS One. 2021; 16: e0245057. https://doi.org/10.1371/journal.pone.0245057

21. van der Velden PG, Marchand M, Cuelenaere B, Das M. Pre-outbreak determinants of perceived risks of corona infection and preventive measures taken. A prospective population-based study. PLoS One. 2020; 15: e0234600. https://doi.org/10.1371/journal.pone.0234600

22. Modini M, Joyce S, Mykletun A, Christensen H, Bryant RA, Mitchell PB, Harvey SB. The mental health benefits of employment: Results of a systematic meta-review. Australas Psychiatry. 2016; 24: 331-633. https://doi.org/10.1177/1039856215618523

23. Read JR, Sharpe L, Modini M, Dear BF. Multimorbidity and depression: A systematic review and meta-analysis. J Affect Disord. 2017; 221: 36-46. https://doi.org/10.1016/j.jad.2017.06.009

24. Roberts T, Miguel Esponda G, Krupchanka D, Shidhaye R, Patel V, Rathod S. Factors associated with health service utilisation for common mental disorders: a systematic review. BMC Psychiatry. 2018; 18-262: 1-19. https://doi.org/10.1186/s12888-018-1837-1

25. Steel Z, Marnane C, Iranpour C, Chey T, Jackson JW, Patel V, Silove D. The global prevalence of common mental disorders: a systematic review and meta-analysis 1980- 2013. Int J Epidemiol. 2014; 43: 476-493. https://doi.org/10.1093/ije/dyu038

26. CBS (Statistics Netherlands). Almost 169 thousand people died in 2020, 10 percent more than expected (in Dutch) https://www.cbs.nl/nl-nl/nieuws/2021/04/bijna-169-duizend-mensen-overleden-in-2020-10-procent-meer-dan-verwacht (accessed February 10, 2021).

27. Scherpenzeel A, Das M. True longitudinal and probability-based internet panels: evidence from The Netherlands. In: Das M, Ester P, Kaczmirek L, editors. Social and behavioral research and the internet: advances in applied methods and research strategies. Taylor & Francis, New York, 2011, 77–104. https://doi.org/10.4324/9780203844922

28. Ware JE, Sherbourne CD. The MOS 36-item short-form health survey, SF-36: I. Conceptual framework and item selection. Med. Care 1992; 30; 473-483. https://doi.org/10.2307/3765916

29. Means-Christensen AJ, Arnau RC, Tonidandel AM, Bramson R, Meagher MW. An efficient method of identifying major depression and panic disorder in primary care. J Behav Med. 2005; 28: 565-572. https://doi.org/10.1007/s10865-005-9023-6

30. Driessen M. Een beschrijving van de MHI-5 in de gezondheidsmodule van het Permanent Onderzoek Leefsituatie [A Description of the MHI-5 in the Health Module of Permanent Research of Living Conditions, POLS]. Den Haag, Statistics Netherlands, 2011.

31. Ormel J, VonKorff M, Ustun TB, Pini S, Korten A, Oldehinkel T. Common mental disorders and disability across cultures. Results from the WHO Collaborative Study on Psychological Problems in General Health Care. JAMA. 1994; 272:1741-1748. https://doi.org/10.1001/jama.272.22.1741

32. Eurostat, 2016. European Health Interview Survey. http://ec.europa.eu/eurostat/documents/3859598/5926729/KS-RA-13-018-EN.PDF/26c7ea80-01d8-420ebdc6-e9d5f6578e7c (Accessed February 2, 2016)

33. de Graaf R, ten Have M, van Gool C, van Dorsselaer S. Prevalence of mental disorders and trends from 1996 to 2009. Results from the Netherlands Mental Health Survey and Incidence Study-2. Soc Psychiatry Psychiatr Epidemiol. 2012; 47: 203-213. https://doi.org/10.1007/s00127-010-0334-8

34. Hyland P, Shevlin M, McBride O, Murphy J, Karatzias T, Bentall RP, et al. Anxiety and depression in the Republic of Ireland during the COVID-19 pandemic. Acta Psychiatr Scand. 2020; 142: 249-256. https://doi.org/10.1111/acps.13219

35. Heinrich LM, Gullone E. The clinical significance of loneliness: a literature review. Clin Psychol Rev. 2006; 26: 695-718. https://doi.org/10.1016/j.cpr.2006.04.002

36. Leigh-Hunt N, Bagguley D, Bash K, Turner V, Turnbull S, Valtorta N, Caan W. An overview of systematic reviews on the public health consequences of social isolation and loneliness. Public Health, 2017; 152:157-171. https://doi.org/10.1016/j.puhe.2017.07.035

37. McKee-Ryan F, Song Z, Wanberg CR, Kinicki AJ. Psychological and physical well-being during unemployment: a meta-analytic study. J Appl Psychol. 2005; 90: 53-76. https://doi.org/10.1037/0021-9010.90.1.53

38. OECD. Unemployment rate (indicator). https://doi.org/10.1787/52570002-en. (accessed February 7, 2021).

39. CBS. Health in Corona times (In Dutch), The Hague, Statistics Netherlands. https://www.cbs.nl/nl-nl/visualisaties/welvaart-in-coronatijd/gezondheid-in-coronatijd (accessed February 10, 2021).

40. de Vries M, Claassen L, te Wierik MJM, van den Hof S, Brabers AEM, de Jong JD, Timmermans DRM, Timen A. Dynamic public perceptions of the coronavirus disease crisis, the Netherlands, 2020. Emerg Infect Dis. 2021 (Early release). https://doi.org/10.3201/eid2704.203328

41. Demyttenaere K, Bruffaerts R, Posada-Villa J, Gasquet I, Kovess V, Lepine JP, Angermeyer MC, Bernert S, de Girolamo G, Morosini P, Polidori G, Kikkawa T, Kawakami N, Ono Y, Takeshima T, Uda H, Karam EG, Fayyad JA, Karam AN, Mneimneh ZN, Medina-Mora ME, Borges G, Lara C, de Graaf R, Ormel J, Gureje O, Shen Y, Huang Y, Zhang M, Alonso J, Haro JM, Vilagut G, Bromet EJ, Gluzman S, Webb C, Kessler RC, Merikangas KR, Anthony JC, Von Korff MR, Wang PS, Brugha TS, Aguilar-Gaxiola S, Lee S, Heeringa S, Pennell BE, Zaslavsky AM, Ustun TB, Chatterji S; WHO World Mental Health Survey Consortium. Prevalence, severity, and unmet need for treatment of mental disorders in the World Health Organization World Mental Health Surveys. JAMA. 2004; 291: 2581-2590. https://doi.org/10.1001/jama.291.21.2581

42. Hentschel S, Eid M, Kutscher T. The influence of major life events and personality traits on the stability of affective well-being. J Happiness Stud. 2016; 18: 719-741. https://doi.org/10.1007/s10902-016-9744-y

43. van der Velden PG, Komproe I, Contino C, de Bruijne M, Kleber RJ, Das M, Schut H. Which groups affected by Potentially Traumatic Events (PTEs) are most at risk for a lack of social support? A prospective population-based study on the 12-month prevalence of PTEs and risk factors for a lack of post-event social support. PLoS One. 2020; 15: e0232477. https://doi.org/10.1371/journal.pone.0232477

44. Harkness KL, Hayden EP, Lopez-Duran NL. Stress sensitivity and stress sensitization in psychopathology: an introduction to the special section. J Abnorm Psychol. 2015; 124:1- 3. https://doi.org/10.1037/abn0000041

45. Sheridan Rains L, Johnson S, Barnett P, Steare T, Needle JJ, Carr S, et al., COVID-19 Mental Health Policy Research Unit Group, 2020. Early impacts of the COVID-19 pandemic on mental health care and on people with mental health conditions: framework synthesis of international experiences and responses. Soc Psychiatry Psychiatr Epidemiol. 2021; 56: 13-24. https://doi.org/10.1007/s00127-020-1924-7

