## appendix 1 for "The prevalence, incidence and risk factors of mental health problems and mental health services use before and 9 months after the COVID-19 outbreak among the general Dutch population. A 3-wave prospective study"

**S1 Appendix Age categories**

|  | 2018 | 2019 | 2020 | T1 vs. T3 | T2 vs. T3 | T1 vs. T2 |
| --- | --- | --- | --- | --- | --- | --- |
| **Prevalence** | n (%) | n (%) | n (%) | aOR (956% CI) | aOR (956% CI) | aOR (956% CI) |
| Anxiety and depression symptoms^1^ | | | | | | |
| - 18-34 years | 216 (19.9) | 224 (20.7) | 242 (22.3) | 1.30 (1.08-1.57)^**^ | 1.17 (0.98-1.41) | 1.11 (0.92-1.34) |
| - 35-49 years | 185 (19.3) | 187 (19.4) | 199 (20.7) | 1.10 (0.91-1.32) | 1.08 (0.91-1.28) | 1.02 (0.85-1.22) |
| - 50-64 years | 154 (14.6) | 159 (15.1) | 140 (13.3) | 0.82 (0.68-0.99)^*^ | 0.81 (0.68-0.95)^*^ | 1.02 (0.87-1.21) |
| - ≥ 65 years | 105 (10.9) | 117 (12.1) | 104 (10.7) | 0.95 (0.78-1.16) | 0.88 (0.73-1.05) | 1.09 (0.91-1.30) |
| Sleep problems | | | | | | |
| - 18-34 years | 143 (13.2) | 160 (14.8) | 158 (14.6) | 1.19 (1.02-1.39)^*^ | 1.01 (0.91-1.11) | 1.19 (1.03-1.36)^*^ |
| - 35-49 years | 181 (18.8) | 185 (19.3) | 194 (20.2) | 1.10 (0.96-1.26) | 1.06 (0.96-1.18) | 1.04 (0.94-1.15) |
| - 50-64 years | 273 (26.0) | 290 (27.5) | 282 (26.8) | 1.02 (0.93-1.12) | 0.94 (0.88-1.02) | 1.08 (1.01-1.16)^*^ |
| - ≥ 65 years | 237 (24.5) | 237 (24.5) | 235 (24.3) | 0.98 (0.90-1.06) | 0.99 (0.93-1.06) | 0.99 (0.93-1.05) |
| Fatigue | | | | | | |
| - 18-34 years | 339 (31.3) | 352 (32.5) | 349 (32.2) | 1.08 (0.96-1.20) | 1.00 (0.93-1.08) | 1.08 (0.98-1.19) |
| - 35-49 years | 321 (33.4) | 314 (32.7) | 305 (31.7) | 0.92 (0.84-1.01) | 0.95 (0.89-1.03) | 0.97 (0.91-1.03) |
| - 50-64 years | 343 (32.6) | 344 (32.7) | 335 (31.8) | 0.95 (0.88-1.02) | 0.95 (0.89-1.01) | 1.00 (0.94-1.05) |
| - ≥ 65 years | 271 (28.0) | 283 (29.2) | 278 (28.7) | 1.03 (0.96-1.11) | 0.99 (0.94-1.05) | 1.04 (0.99-1.09) |
| Disabilities due to health problems | | | | | | |
| - 18-34 years | 96 (8.9) | 80 (7.4) | 77 (7.1) | 0.84 (0.63-1.13) | 1.03 (0.71-1.48) | 0.82 (0.59-1.15) |
| - 35-49 years | 97 (10.1) | 86 (9.0) | 80 (8.3) | 0.79 (0.58-1.08) | 0.91 (0.66-1.25) | 0.87 (0.65-1.17) |
| - 50-64 years | 128 (12.2) | 105 (10.0) | 112 (10.6) | 0.78 (0.62-0.99)^*^ | 1.02 (0.81-1.29) | 0.77 (0.62-0.96)^*^ |
| - ≥ 65 years | 83 (8.6) | 74 (7.6) | 78 (8.1) | 0.88 (0.70-1.11) | 1.05 (0.84-1.30) | 0.84 (0.67-1.06) |
| Medicines for anxiety/depression | | | | | | |
| - 18-34 years | 33 (3.0) | 31 (2.9) | 37 (3.4) | 1.17 (0.86-1.59) | 1.21 (1.01-1.46)^*^ | 0.96 (0.73-1.28) |
| - 35-49 years | 61 (6.3) | 58 (6.0) | 58 (6.0) | 0.94 (0.81-1.09) | 1.00 (0.91-1.11) | 0.94 (0.84-1.06) |
| - 50-64 years | 58 (5.5) | 59 (5.6) | 60 (5.7) | 0.99 (0.85-1.15) | 1.00 (0.89-1.13) | 0.99 (0.88-1.10) |
| - ≥ 65 years | 38 (3.9) | 39 (4.0) | 39 (4.0) | 1.04 (0.90-1.19) | 1.02 (0.92-1.13) | 1.02 (0.91-1.14) |
| Medicines for sleep problems | | | | | | |
| - 18-34 years | 21 (1.9) | 39 (4.0) | 29 (2.7) | 1.51 (0.82-2.77) | 1.09 (0.62-1.90) | 1.39 (0.77-2.50) |
| - 35-49 years | 38 (4.0) | 40 (4.2) | 38 (4.0) | 0.97 (0.74-1.28) | 0.95 (0.75-1.20) | 1.03 (0.82-1.29) |
| - 50-64 years | 64 (6.1) | 65 (6.2) | 71 (6.7) | 1.04 (0.86-1.25) | 1.06 (0.92-1.23) | 0.97 (0.84-1.14) |
| - ≥ 65 years | 66 (6.8) | 71 (7.3) | 70 (7.2) | 1.02 (0.86-1.20) | 0.96 (0.84-1.08) | 1.06 (0.94-1.21) |
| Use of mental health services^2^ | | | | | | |
| - 18-34 years | 127 (11.7) | 129 (11.9) | 121 (11.2) | 1.00 (0.77-1.30) | 0.95 (0.74-1.22) | 1.05 (0.85-1.31) |
| - 35-49 years | 101 (10.5) | 100 (10.4) | 101 (10.5) | 0.99 (0.76-1.28) | 1.01 (0.81-1.25) | 0.98 (0.77-1.24) |
| - 50-64 years | 79 (7.5) | 86 (8.2) | 74 (7.0) | 0.89 (0.68-1.16) | 0.83 (0.66-1.05) | 1.07 (0.84-1.35) |
| - ≥ 65 years | 34 (3.5) | 30 (3.1) | 21 (2.2) | 0.60 (0.41-0.87)^**^ | 0.69 (0.49-0.97)^*^ | 0.87 (0.63-1.19) |

^1^According cut-off score of ≤ 59 on MHI-5. ^2^Psychiatrist/psychologist/psychotherapist in past 12 months. aOR=Odds ratio adjusted for sex, age, marital status, employment status, education level and disease at T1, T2, and T3. 95% CI=95% confidence interval for aOR. ^*^ p < 0.05, ^**^ p < 0.01
