## appendix 2 for "The prevalence, incidence and risk factors of mental health problems and mental health services use before and 9 months after the COVID-19 outbreak among the general Dutch population. A 3-wave prospective study"

**S2 Appendix risk factors**

Predictors anxiety and depression symptoms at T2 and T3

|  | **Anxiety and depression symptoms** | |
| --- | --- | --- |
| **Predictors previous year** | 2019 (T2) | 2020 (T3) |
|  | aOR (95% CI) | aOR (95% CI) |
| Employed |  |  |
| - no (ref.) | 1 | 1 |
| - yes | 0.59 (0.47-0.74)*** | 0.76 (0.59-0.96)* |
| Married |  |  |
| - yes (ref.) | 1 | 1 |
| - no | 1.35 (1.09-1.66)** | 1.27 (1.03-1.57)* |
| Education level |  |  |
| - low education (ref.) | 1 | 1 |
| - medium education level | 0.74 (0.58-0.96)* | 0.79 (0.61-1.03) |
| - high education level | 0.76 (0.59-0.98)* | 0.67 (0.52-0.88)** |
| Sex |  |  |
| - male (ref.) | 1 | 1 |
| - female | 1.18 (0.97-1.44) | 0.93 (0.77-1.14) |
| Physical disease |  |  |
| - no (ref.) | 1 | 1 |
| - yes | 1.51 (1.17-1.94)** | 1.76 (1.37-2.28)*** |
| Age category |  |  |
| - 65+ (ref.) | 1 | 1 |
| - 50-64 years | 1.77 (1.29-2.44)*** | 1.73 (1.24-2.41)** |
| - 35-49 years | 2.58 (1.83-3.64)*** | 3.25 (2.27-4.66)*** |
| - 18-34 years | 2.26 (1.62-3.14)*** | 3.19 (2.26-4.51)*** |
| Anxiety and depression sympt. |  |  |
| - no (ref.) | 1 | 1 |
| - yes | 13.36 (10.93-16.34)*** | 14.31 (11.68-17.53)*** |

Predictors sleep problems at T2 and T3

|  | **Sleep problems** | |
| --- | --- | --- |
| **Predictors previous year** | 2019 (T2) | 2020 (T3) |
|  | aOR (95% CI) | aOR (95% CI) |
| Employed | 1 | 1 |
| - no (ref.) | 0.86 (0.61-1.23) | 0.76 (0.52-1.10) |
| - yes |  |  |
| Married | 1 | 1 |
| - yes (ref.) | 0.99 (0.72-1.34) | 1.14 (0.83-1.56) |
| - no |  |  |
| Education level | 1 | 1 |
| - low education (ref.) | 0.83 (0.57-1.21) | 0.97 (0.65-1.44) |
| - medium education level | 0.83 (0.57-1.22) | 1.09 (0.73-1.63) |
| - high education level |  |  |
| Sex | 1 | 1 |
| - male (ref.) | 1.48 (1.11-1.98)** | 1.29 (0.95-1.75) |
| - female |  |  |
| Physical disease | 1 | 1 |
| - no (ref.) | 1.66 (1.16-2.38)** | 2.14 (1.47-3.09)*** |
| - yes |  |  |
| Age category | 1 | 1 |
| - 65+ (ref.) | 1.79 (1.15-2.80)* | 1.64 (1.04-2.60)* |
| - 50-64 years | 1.30 (0.79-2.14) | 1.72 (1.02-2.90)* |
| - 35-49 years | 1.35 (0.83-2.21) | 1.10 (0.66-1.86) |
| - 18-34 years |  |  |
| Sleep problems |  |  |
| - no (ref.) | 1 | 1 |
| - yes | 214.07 (159.01-288.19)*** | 243.35 (179.97-329.04)*** |

Predictors fatigue at T2 and T3

|  | **Fatique** | |
| --- | --- | --- |
| **Predictors previous year** | 2019 (T2) | 2020 (T3) |
|  | aOR (95% CI) | aOR (95% CI) |
| Employed |  |  |
| - no (ref.) | 1 | 1 |
| - yes | 0.76 (0.53-1.08) | 0.87 (0.61-1.25) |
| Married |  |  |
| - yes (ref.) | 1 | 1 |
| - no | 1.11 (0.81-1.50) | 1.26 (0.93-1.70) |
| Education level |  |  |
| - low education (ref.) | 1 | 1 |
| - medium education level | 0.89 (0.61-1.29) | 0.97 (0.66-1.42) |
| - high education level | 0.88 (0.60-1.30) | 0.73 (0.50-1.07) |
| Sex |  |  |
| - male (ref.) | 1 | 1 |
| - female | 1.17 (0.88-1.56) | 1.07 (0.80-1.42) |
| Physical disease |  |  |
| - no (ref.) | 1 | 1 |
| - yes | 1.76 (1.21-2.55)** | 1.93 (1.34-2.78)*** |
| Age category |  |  |
| - 65+ (ref.) | 1 | 1 |
| - 50-64 years | 1.18 (0.75-1.87) | 1.38 (0.87-2.17) |
| - 35-49 years | 1.17 (0.71-1.93) | 1.43 (0.86-2.36) |
| - 18-34 years | 1.50 (0.93-2.42) | 1.46 (0.90-2.37) |
| Fatigue |  |  |
| - no (ref.) | 1 | 1 |
| - yes | 276.37 (207.38-368.31)*** | 263.58 (198.27-350.40)*** |

Predictors disabilities at T2 and T3

|  | **Disabilities** **due to health problems** | |
| --- | --- | --- |
| **Predictors previous year** | 2019 (T2) | 2020 (T3) |
|  | aOR (95% CI) | aOR (95% CI) |
| Employed | 1 | 1 |
| - no (ref.) | 0.60 (0.45-0.81)** | 0.54 (0.40-0.72)*** |
| - yes |  |  |
| Married | 1 | 1 |
| - yes (ref.) | 1.01 (0.78-1.32) | 1.05 (0.81-1.35) |
| - no |  |  |
| Education level | 1 | 1 |
| - low education (ref.) | 0.60 (0.44-0.82)** | 1.16 (0.85-1.58) |
| - medium education level | 0.69 (0.50-0.95)* | 0.99 (0.71-1.36) |
| - high education level |  |  |
| Sex | 1 | 1 |
| - male (ref.) | 1.54 (1.19-1.99)** | 1.14 (0.89-1.46) |
| - female |  |  |
| Physical disease | 1 | 1 |
| - no (ref.) | 2.32 (1.74-3.09)*** | 2.28 (1.73-3.01)*** |
| - yes |  |  |
| Age category | 1 | 1 |
| - 65+ (ref.) | 1.95 (1.34-2.85)** | 1.95 (1.37-2.78)*** |
| - 50-64 years | 2.51 (1.64-3.83)*** | 2.02 (1.34-3.04)** |
| - 35-49 years | 2.00 (1.31-3.07)** | 1.40 (0.93-2.13) |
| - 18-34 years |  |  |
| Disabilities |  |  |
| - no (ref.) | 1 | 1 |
| - yes | 10.99 (8.45-14.31)*** | 8.78 (6.69-11.53)*** |

Predictors mental health services use at T2 and T3

|  | **Mental health services use** | |
| --- | --- | --- |
| **Predictors previous year** | 2019 (T2) | 2020 (T3) |
|  | aOR (95% CI) | aOR (95% CI) |
| Employed |  |  |
| - no (ref.) | 1 | 1 |
| - yes | 0.82 (0.61-1.11) | 0.46 (0.34-0.63)*** |
| Married |  |  |
| - yes (ref.) | 1 | 1 |
| - no | 1.02 (0.77-1.36) | 1.11 (0.83-1.50) |
| Education level |  |  |
| - low education (ref.) | 1 | 1 |
| - medium education level | 0.81 (0.57-1.16) | 1.09 (0.75-1.60) |
| - high education level | 0.98 (0.68-1.40) | 1.18 (0.80-1.74) |
| Sex |  |  |
| - male (ref.) | 1 | 1 |
| - female | 1.59 (1.22-2.07)** | 1.62 (1.22-2.15)** |
| Physical disease |  |  |
| - no (ref.) | 1 | 1 |
| - yes | 1.52 (1.08-2.13)* | 1.63 (1.15-2.31)** |
| Age category |  |  |
| - 65+ (ref.) | 1 | 1 |
| - 50-64 years | 2.91 (1.78-4.76)*** | 4.13 (2.43-7.01)*** |
| - 35-49 years | 3.67 (2.18-6.17)*** | 7.52 (4.35-13.01)*** |
| - 18-34 years | 4.01 (2.41-6.66)*** | 5.77 (3.35-9.92)*** |
| Mental health services use |  |  |
| - no (ref.) | 1 | 1 |
| - yes | 22.53 (17.13-29.63)*** | 21.98 (16.56-29.16)*** |
